# The Association between Afghan Women’s Autonomy and Experience of Domestic Violence, Moderated by Education Status

**DOI:** 10.1101/2024.04.28.24305784

**Authors:** Sahra Ibrahimi, Marie E. Thoma

## Abstract

This study examines the association between Afghan women’s autonomy (WA) and experience of domestic violence (physical, sexual, and emotional) in the previous 12 months, and whether this association is moderated by education status. We used data from 19,098 married women aged 15-49, who completed the 2015 Afghanistan Demographic and Health Survey- the first and only national survey administered in the country. WA was measured across 5 domains (healthcare, visiting family, household purchases, spending, and contraceptive use). Adjusted odds ratios and 95% confidence intervals for the association between domestic violence in the past 12 months (any vs. none) and WA were estimated using multiple logistic regression and adjusted for covariates. Interaction terms between education status and WA were also assessed. We found that the experience of physical, emotional, and sexual violence was 45% 30%, and 7%, and at least 1 in 2 had no autonomy. After adjustment, compared to women without autonomy, WA in healthcare decisions, spending, visiting families, and household purchases significantly decreased the odds of physical violence. Similarly, WA in healthcare decisions and spending significantly decreased the odds of sexual violence. Lastly, WA in spending and not using contraception was associated with reduced odds of emotional violence. We also found a greater protective effect of WA in visiting family among women with any education across each domestic violence outcome. These findings provide insights into areas for intervention to address gender inequalities (Sustainable Development Goal 3) and mitigate adverse health outcomes for mothers and their children (Goal 5).

## Introduction

Women’s autonomy has been defined as the ability of women to access information and use it in making decisions about their lives and environments.^1,2^ The prevalence of gender inequalities and lack of women’s autonomy in the world is high. A report by the United Nations shows that globally only 57% of women make decisions about sexual relations, contraceptive use, and reproductive health care.^3^ Lack of women’s autonomy is especially worse in low-income countries and patriarchal cultures. In Afghanistan, only 5% of women make decisions alone about their health care, and 44% of women’s health care decisions are made by their husbands.^4^ In general, women’s autonomy facilitates access by women to social resources including education, power, and prestige,^5,6^ and material resources including job, land, wealth,^5,7^ and healthcare resources.^8–10^ In return, greater access to resources can reduce gender inequities and improve women’s quality of life, including reducing experiences of domestic violence^11^ and adverse health outcomes.^10,12^ One of the United Nations’ top priorities is reducing gender inequalities and violence against women through the Sustainable Development Goal 5 (SDG 5).^13^ Overall, SDG5 seeks to promote gender equality and empower women and girls to fully participate in all aspects of society, which has shown to have positive impacts on health.^14^ Therefore, promoting gender equality and women’s autonomy also help with achieving SDG 3, which “ensure healthy lives and promote well-being for all at all ages.”^15^

Studies show that higher women’s autonomy is associated with increased use of contraception,^16,17^ reduced experience of domestic violence,^11,18^ decreased unwanted pregnancy,^19^ negative attitudes toward female genital cutting,^20^ and decreased maternal, infant and child mortality.^21–25^ However, these findings are not always consistent across geographic regions or contexts with some studies showing women’s autonomy to be associated with increased experience of violence^26,27^ and increased unintended pregnancy.^28^ These inconsistent findings exist because the degree and direction of the association between women’s autonomy and health outcomes differ across countries based on their laws, social norm, and cultural context.^29^ For example, while, studies in Bangladesh^18^ and Ghana^11^ found a significant association between women’s autonomy and decreased experience of intimate partner violence, two other studies in the Philippines^26^ and India^27^ found women’s autonomy to be associated with increased experience of domestic violence. The authors of the two studies in the Philippines and India attributed their findings to the fact that when traditional gender norms of men being the dominant decision-makers are challenged, men may become violent to restore their dominance.^27^

Women’s autonomy and domestic violence are persistent issues in Afghanistan. The presence of war and political instability in the nation since 1973 has resulted in the emergence of structural violence.^30,31^ This type of violence is characterized as an indirect form of aggression that is deeply embedded within oppressive social systems, potentially influencing people’s self-perceptions, actions, and customs.^31^ War deprives people of educational and employment opportunities, which often results in poverty, inequality, oppression, and stress.^32^ These factors contribute to an increase in domestic violence and the reinforcement of patriarchy. Prior studies have suggested that men, who have fewer opportunities for employment and supporting their families, may suffer from a sense of inadequacy as providers and experience stress and anxiety that may be expressed in the form of violence.^32^

Given potential contextual differences in associations between women’s autonomy and experience of domestic violence, it is important to examine this subject in Afghanistan, specifically, given Afghanistan’s unique political and sociocultural environment that shape its population health. Afghanistan has the highest unequal gender norms and prevalence of domestic violence (56%) in the world.^4,33^ This has worsened with recent conflicts. The 2021 overtake of the country by the Taliban has exacerbated the violation of women’s rights in Afghanistan.^34^ The Taliban has removed the Elimination of Violence against Women (EVAW Law) by the United Nations Assistance Mission in Afghanistan (UNAMA).^34^ The country’s Women, Peace, and Security (WPS) Index has decreased by 28% since 2017.^35^ The Taliban has also eliminated the Ministry of Women’s Affairs, which protected women’s rights by establishing and enforcing policies.^34^ Furthermore, women are currently banned from getting an education beyond primary school or participating in government; women are not even allowed to step outside their homes without being accompanied by a male relative.^36^ These policies limit women’s autonomy and can potentially exacerbate domestic violence.

Despite the alarming prevalence of reduced women’s autonomy in Afghanistan and high rates of domestic violence, there is no research linking women’s autotomy to domestic violence in Afghanistan. Also, no previous study has looked at the moderating effect of education on this association. A study in Ethiopia showed that women who had primary and secondary or above education were about two and four times more likely to have greater autonomy than women with no education.^37^ Similarly, a study using a national representative sample from Bangladesh found that women with greater autonomy were more likely to have higher education level.^38^ On another hand, education is inversely associated with domestic violence. Studies show that women’s higher levels of education are associated with lower levels of domestic violence.^39^ Specifically, a study in Ghana found that compared to women with no education, women with some education were less likely to experience physical or emotional violence from their partners.^11^ Depending on the status of women’s education, the effect of women’s autonomy on their experience of domestic violence may differ. Accordingly, this study aims to examine the association between women’s autonomy and experience of domestic violence (physical, sexual, and emotional) in the past 12 months, and whether this association is moderated by education status. We hypothesize that there will be a significant increase in odds of domestic violence among women who lack autonomy compared to women who don’t. We also hypothesize that women with some education will have a lower magnitude of association between women’s autonomy and experience of domestic violence as compared to women with no education.

## Methods

### Data Sources and Sampling

We examined data from the 2015 Afghanistan Demographic and Health Survey (AfDHS), which is the first and only nationally conducted cross-sectional survey in Afghanistan.^4^ The survey program, Demographic and Health Surveys (DHS), is supported by the United States Agency for International Development (USAID), and operates in 90 nations.^40^ Due to the continuous war in Afghanistan, AfDHS was conducted only once in 2015. The 2015 AfDHS collected data from all 34 provinces of the country, interviewing Afghan civilian non-institutionalized population (women and men) ages 15-49, and provided up-to-date data on several subjects.^4^ In this study, we used modules on women’s empowerment, domestic violence, infant and child mortality, and demographics that include the variables of interest. The data was obtained from DHS’s official website.

AfDHS data collection was conducted through interviews from June 15, 2015, to February 23, 2016.^4^ AfDHS questionnaires were pre-tested before the fieldwork to test the instrument validity and maintain data quality. Additionally, to reduce data entry errors, the CSPro software was used for data entry, and double entry was done for complete verification.^4^ A two-stage stratified sampling approach was adopted, with sampling weights determined based on the likelihood of sampling, which was computed for each cluster and sampling phase. In the initial stage, 950 clusters containing enumeration areas were identified and chosen.^4^ In the second stage, about 27 households per cluster were systematically sampled, resulting in 25,650 households, out of which 24,395 were interviewed (accounting for a response rate of 98%).^4^ Heeding the World Health Organization (WHO) ethical guidelines, only one woman per household (a total of 21,342 women) were selected to answer the questions in the domestic violence module. The detailed methodology for AfDHS data can be found in AfDHS 2015 final report.^4^

In this study, the sample was restricted to currently married women (n=19,089), aged 15-49, who completed the domestic violence module. Since sex before marriage is illegal in the Islamic Republic of Afghanistan, AfDHS could only ask married individuals about reproductive and perinatal health outcomes.^4^ The Inner-City Fund (ICF) Institutional Review Board (IRB) and Afghanistan’s Ministry of Public Health approved data collection tools and procedures.^4^ We also received IRB exemption from the University of Maryland IRB in College Park.

### Measurement and Operationalization of Variables

Women’s autonomy was measured by asking women if they usually made decisions a) alone, b) jointly with their husband, c) their husband alone, or d) someone else/other in the following five areas: 1) the woman’s health care, 2) contraceptive use, 3) major household purchases, 4) how to use women’s earnings, and 5) visits to the woman’s family or relatives.^4^ The four response options for each of the five women’s autonomy items were grouped based on the existing literature;^8^ response options (a) and (b) were coded as women’s autonomy, and response options (d) and (c) were coded as women’s lack of autonomy. For contraceptive use, women who did not use contraception were not asked the question on decision-making. We, therefore, included a third category for this dimension of WA to include “not using,” which allowed us to include information from all women in the analytic sample.

In this study, we considered all three types of domestic violence: verbal, physical, and sexual violence in the previous 12 months. For physical domestic violence, women were asked if, “in the past 12 months, any household members (i.e., current husband, former husband, mother/stepmother, father/stepfather, mother-in-law, father-in-law, or siblings) push you, shake you, or throw something at you; slap you; twist your arm or pull your hair; punch you with his fist or with something that could hurt you; kick you, drag you, or beat you up; try to choke you or burn you on purpose; or threaten or attack you with a knife, gun, or any other weapon.”^4^ For sexual domestic violence, women were asked if, “in the past 12 months any household members physically force you to have sexual intercourse with them even when you did not want to; physically force you to perform any other sexual acts you did not want to; force you with threats or any other way to perform sexual acts that you did not want to do.”^4^ For emotional violence, women were asked if, “in the past 12 months, any household members say or do something to humiliate you in front of others; threaten to hurt or harm you or someone close to you; insult you or make you feel bad about yourself.”^4^ The binary variables mentioned above were classified as “yes” if a woman reported enduring any type of violence within each respective category.

Education was assessed as a possible moderator of the association between women’s autonomy and domestic violence. Educational attainment was measured by asking women if they ever attended school, and if yes, what was the number of years of schooling completed.^4^ We grouped education into no education (never attended school), and some education (primary, secondary, or higher). We grouped primary, secondary, or higher education into one group because our preliminary analysis showed very little difference in moderation by primary vs. secondary or higher education groups.

We also considered the following additional covariates as confounders in regression models: age, ethnicity, wealth, residency (urban vs. rural), and parity. We controlled for these variables because based on the literature review^11,18^ and our analysis, these variables were associated with both WA and experience of domestic violence. Age was categorized as 15–24, 25–34, and 35–49. Ethnicity included Pashtun, Tajik, Hazara, and others (Uzbek, Turkmen, Nurstani, Baloch, and Pashai). The wealth index was classified into rich, middle, and poor, and the place of residence was dichotomized as urban and rural. Parity was grouped as 4 children or less or 5 or more children.

### Analysis

First, we examined the distribution of the outcome and covariates by whether women experienced autonomy across each of the five WA domains. Chi-squared tests were used to examine differences in the distribution of each covariate and domestic violence outcome for women who experience autonomy vs. those who did not in each WA domain.

Multiple logistic regression models were used to examine the association between women’s autonomy categories and types of experience of domestic violence in the past 12 months. The results are presented as adjusted odds ratios (ORs) with 95% confidence intervals, and all tests of hypothesis are 2-tailed with a type 1 error rate set at 5%. In Model 1, we presented unadjusted ORs, in Model 2, we adjusted for sociodemographic, and in Model 3, we adjusted for both sociodemographic and women’s autonomy variables. We assessed multi-collinearity using variance inflation factors (VIF).^41^ Additionally, the moderating effect of education on the association between women’s autonomy categories and types of domestic violence was explored using interaction terms between education (any vs. none) and each WA variable separately. Associations between WA and each domestic violence outcome were stratified by educational attainment.

Analyses were conducted using Stata 17, and weighted for the complex survey design.

## Results

In our sample of 19,098 Afghan women, 45% (n=8,525) of women experienced physical violence, 30% (5,671) experienced emotional violence, and 7% (1,411) experienced sexual violence. Women’s autonomy was 49% (4,324) in healthcare decision making, 38% (6,939) in spending, 47% (8,894) in household purchases, 58% (11,005) in visiting families, and 19.9% (3,749) in contraception use. In the bivariate analysis (**Table 1**), each of the women’s autonomy categories was associated with types of domestic violence. Compared with those women with no autonomy in healthcare decision making, those with autonomy were less likely to experience physical violence (54.0% vs. 43.3%, p < 0.000), sexual violence (8.1% vs. 4.6%, p < 0.000), and emotional violence (54% vs. 43%, p < 0.000). Similarly, those with household purchase autonomy were less likely to experience physical violence (54.4% vs. 41.4%, p < 0.000), sexual violence (7.5% vs. 4.9%, p < 0.000), and emotional violence (39.9% vs. 32.3%, p < 0.000). There were also significant differences in experience of domestic violence among women with autonomy in visiting families vs. women with no autonomy. Those with autonomy in visiting families were less likely to experience physical violence (54.3% vs. 44.2%, p < 0.000), sexual violence (6.9% vs. 6.0%, p < 0.000), and emotional violence (39.4% vs. 33.3%, p < 0.000). In general, women with autonomy were more likely to be older, Tajik, middle class, and live in urban areas.

**Table 1:** Bivariate analysis between women’s autonomy and selected characteristics of married women ages 15-49, Afghanistan DHS 2015.

| Variables | Total<br>N (%) | Healthcare<br>n (%) |  | Spending<br>n (%) |  | Household<br>purchases n (%) |  | Visiting families<br>n (%) |  | Contraception<br>n (%) |  |  |
| --- | --- | --- | --- | --- | --- | --- | --- | --- | --- | --- | --- | --- |
|  | 19,098 | No | Yes | No | Yes | No | Yes | No | Yes | No | Yes | Not using |
|  |  | 9,758 | 9,324 | 11,506 | 6,939 | 10,184 | 8,894 | 8,076 | 11,005 | 489 | 3,749 | 14,641 |
| **Physical** |  |  |  |  |  |  |  |  |  |  |  |  |
| No | 10,573<br>(55.4) | 4844<br>(46.0) | 5722<br>(56.7) | 5678<br>(46.4) | 4480<br>(59.7) | 5081<br>(45.6) | 5479<br>(58.6) | 4091<br>(45.7) | 6473<br>(55.8) | 210<br>(43.5) | 1935<br>(45.6) | 8319<br>(53.2) |
| Yes | 8,525<br>(44.6) | 4914<br>(54.0) | 3602<br>(43.3) | 5828<br>(53.6) | 2459<br>(40.4) | 5103<br>(54.4) | 3415<br>(41.4) | 3985<br>(54.3) | 4532<br>(44.2) | 279<br>(56.5) | 1814<br>(54.4) | 6322<br>(46.8) |
| **Sexual** |  |  |  |  |  |  |  |  |  |  |  |  |
| No | 17,687<br>(92.6) | 8778<br>(91.9) | 8893<br>(95.5) | 10428<br>(92.8) | 6638<br>(94.8) | 9216<br>(92.5) | 8451<br>(95.1) | 7409<br>(93.1) | 10261<sup>ns</sup><br>(94.0) | 459<br>(93.9) | 3411<br>(92.8) | 13618<sup>ns</sup><br>(93.8) |
| Yes | 1,411<br>(7.4) | 980<br>(8.1) | 431<br>(4.6) | 1078<br>(7.2) | 301<br>(5.2) | 968<br>(7.5) | 443<br>(4.9) | 667<br>(6.9) | 744<br>(6.0) | 30<br>(6.1) | 338<br>(7.2) | 1023<br>(6.2) |
| **Emotional** |  |  |  |  |  |  |  |  |  |  |  |  |
| No | 13,427<br>(70.3) | 6550<br>(61.1) | 6867<br>(66.9) | 7637<br>(60.3) | 5314<br>(70.5) | 6857<br>(61.1) | 5520<br>(67.7) | 7897<br>(60.6) | 261<br>(66.7) | 2403<br>(52.2) | 10616<br>(55.5) | 6550<br>(66.8) |
| Yes | 5,671<br>(29.7) | 3208<br>(38.9) | 2457<br>(33.1) | 3869<br>(39.7) | 1625<br>(29.5) | 3327<br>(38.9) | 2556<br>(32.3) | 3108<br>(39.4) | 228<br>(33.3) | 1346<br>(47.8) | 4025<br>(44.6) | 3208<br>(33.2) |
| **Age** |  |  |  |  |  |  |  |  |  |  |  |  |
| 15-24 | 3,739<br>(19.6) | 2,142<br>(24.8) | 1,591<br>(19.8) | 2,447<br>(23.7) | 1,155<br>(19.9) | 2,289<br>(25.3) | 1,446<br>(18.4) | 1,983<br>(28.3) | 1,752<br>(17.4) | 94<br>(21.3) | 533<br>(18.1) | 3,075<br>(23.6) |
| 25-34 | 7,750<br>(40.6) | 4,174<br>(40.2) | 3,570<br>(36.6) | 4,927<br>(39.4) | 2,655<br>(37.6) | 4,307<br>(39.3) | 3,431<br>(37.2) | 3,440<br>(39.2) | 4,303<br>(37.8) | 213<br>(36.0) | 1,470<br>(36.2) | 5,996<br>(39.4) |
| 35-49 | 7,609<br>(39.8) | 3,442<br>(35.1) | 4,163<br>(43.6) | 4,132<br>(36.8) | 3,129<br>(42.5) | 3,588<br>(35.3) | 4,017<br>(44.4) | 2,653<br>(32.5) | 4,950<br>(44.8) | 182<br>(42.6) | 1,746<br>(45.7) | 5,570<br>(37.0) |
| **Education** |  |  |  |  |  |  |  |  |  |  |  |  |
| None | 16,819<br>(86.0) | 8,725<br>(88.8) | 7,664<br>(80.0) | 10,201<br>(87.0) | 5,638<br>(79.3) | 9,006<br>(86.7) | 7,380<br>(81.6) | 7,211<br>(87.0) | 9,177<br>(82.4) | 470<br>(82.4) | 2,985<br>(77.4) | 16,256<br>(86.6) |
| Primary+ | 2,746<br>(14.0) | 1,033<br>(11.2) | 1,660<br>(20.0) | 1,305<br>(13.0) | 1,301<br>(20.7) | 1,178<br>(13.3) | 1,514<br>(18.5) | 865<br>(13.0) | 1,828<br>(17.6) | 77<br>(17.6) | 764<br>(22.6) | 2,681<br>(13.5) |
| **Ethnicity** |  |  |  |  |  |  |  |  |  |  |  |  |
| Pashtun | 7,441<br>(39) | 4,663<br>(50.3) | 2,775<br>(28.4) | 5,127<br>(44.4) | 2,118<br>(31.3) | 4,712<br>(47.0) | 2,725<br>(29.9) | 4,104<br>(53.2) | 3,332<br>(28.5) | 302<br>(50.5) | 1,479<br>(38.5) | 5,570<br>(39.4) |
| Tajik | 6,277<br>(32.9) | 2,821<br>(27.6) | 3,445<br>(37.7) | 3,472<br>(28.7) | 2,609<br>(40.7) | 2,973<br>(27.9) | 3,296<br>(38.7) | 2,167<br>(25.8) | 4,100<br>(38.1) | 173<br>(36.1) | 1,494<br>(41.6) | 4,569<br>(29.9) |
| Hazara | 1,978<br>(10.4) | 672<br>(7.5) | 1,306<br>(11.2) | 845<br>(7.0) | 1,038<br>(13.4) | 663<br>(6.8) | 1,315<br>(12.7) | 472<br>(6.2) | 1,506<br>(11.9) | 43<br>(6.8) | 414<br>(8.6) | 1,502<br>(9.7) |
| Uzbek | 1,442<br>(7.6) | 635<br>(7.8) | 805<br>(15.1) | 902<br>(12.3) | 450<br>(8.4) | 711<br>(10.8) | 728<br>(12.0) | 424<br>(8.1) | 1,016<br>(13.9) | 14<br>(4.0) | 175<br>(7.3) | 1,245<br>(12.7) |
| Others | 1,931<br>(10.1) | 950<br>(6.8) | 981<br>(7.6) | 1,138<br>(7.6) | 718<br>(6.3) | 1,107<br>(7.5) | 819<br>(6.7) | 897<br>(6.7) | 1,034<br>(7.6) | 15<br>(2.6) | 185<br>(4.1) | 1,728<br>(8.3) |
| **Residency** |  |  |  |  |  |  |  |  |  |  |  |  |
| Urban | 4,760<br>(24.9) | 2,218<br>(20.4) | 2,536<br>(24.8) | 2,706<br>(20.9) | 1,900<br>(26.1) | 2,401<sup>ns</sup><br>(21.6) | 2,355<br>(23.8) | 1,811<br>(20.2) | 2,946<br>(24.5) | 162<br>(33.0) | 1,363<br>(36.1) | 3,186<br>(18.3) |
| Rural | 14,338<br>(75.1) | 7,540<br>(79.6) | 6,788<br>(75.2) | 8,800<br>(79.1) | 5,039<br>(73.9) | 7,783<br>(78.4) | 6,539<br>(76.2) | 6,265<br>(79.8) | 8,059<br>(75.5) | 327<br>(67.0) | 2,386<br>(63.9) | 11,455<br>(81.7) |
| **Wealth** |  |  |  |  |  |  |  |  |  |  |  |  |
| Poor | 8,083<br>(42.3) | 3,989<br>(40.6) | 4,090<br>(41.3) | 4,785<br>(40.8) | 2,984<br>(40.7) | 4,116<br>(39.0) | 3,961<br>(43.5) | 3,204<br>(38.6) | 4,874<br>(42.8) | 140<br>(28.6) | 1,173<br>(30.5) | 6,671<br>(44.4) |
| Middle | 4,076<br>(21.4) | 2,289<br>(23.0) | 1,783<br>(17.6) | 2,649<br>(22.2) | 1,294<br>(16.6) | 2,326<br>(22.0) | 1,745<br>(18.3) | 1,959<br>(23.5) | 2,113<br>(17.8) | 98<br>(18.1) | 697<br>(15.6) | 3,230<br>(21.8) |
| Rich | 6,939<br>(36.4) | 3,480<br>(36.4) | 3,451<br>(41.1) | 4,072<br>(37.0) | 2,661<br>(42.8) | 3,742<br>(39.1) | 3,188<br>(38.3) | 2,913<br>(37.9) | 4,018<br>(39.4) | 251<br>(53.3) | 1,879<br>(53.9) | 4,740<br>(33.8) |

| Parity |  |  |  |  |  |  |  |  |  |  |  |  |
| --- | --- | --- | --- | --- | --- | --- | --- | --- | --- | --- | --- | --- |
| <4 | 9,166<br>(48) | 4,956<br>(54.9) | 4,201<br>(49.3) | 5,792<sup>ns</sup><br>(53.4) | 3,096<br>(50.4) | 5,222<br>(55.2) | 3,931<br>(48.1) | 4,288<br>(57.5) | 4,869<br>(47.8) | 256<br>(54.7) | 1,451<br>(42.2) | 7,372<br>(55.1) |
| >=5 | 9,932<br>(52) | 4,802<br>(45.1) | 5,123<br>(50.7) | 5,714<br>(46.6) | 3,843<br>(49.6) | 4,962<br>(44.8) | 4,963<br>(51.9) | 3,788<br>(42.5) | 6,136<br>(52.2) | 233<br>(45.3) | 2,298<br>(57.8) | 7,269<br>(44.9) |

*Footnote:* P-values compare the violence measures and sociodemographic within each women's autonomy measure; all P-values were significant (>0.000) except for the comparisons noted by "ns." All percent values are weighted using svy command. Missing values were only <0.15% for contraception, ethnicity, household purchases, visiting families, and healthcare. Spending had 3.4% missingness.

**Table 2** presents multiple logistic regression models for the association between types of violence and categories of women’s autonomy. In Model 1, we indicate unadjusted odds ratios, while in Model 2, we adjust for relevant confounders including age, ethnicity, wealth, residency, and parity. After adjusting for confounders, women’s autonomy in healthcare decisions (Adjusted odds ratio [AOR]=0.70, CI: 0.60-0.81, *p<0.000*), spending (AOR=0.58, CI: 0.51-0.66, *p<0.000*), visiting families (AOR=0.69, CI: 0.60-0.80, *p<0.000*), household purchases (AOR=0.59, CI: 0.52-0.68, *p<0.000*), and not using contraception (AOR=0.66, CI: 0.46-0.93, *p<0.01*) were significantly associated with decreased experience of physical violence. In addition, women’s autonomy in healthcare decisions (AOR = 0.51, 95% CI: 0.39-0.65, *p<0.000*), spending (AOR=0.62, CI: 0.48-0.80, *p<0.000*), and household purchases (AOR=0.56, CI: 0.43-0.72, *p<0.000*) were significantly associated with decreased experience of sexual violence. Lastly, women’s autonomy in healthcare (AOR=0.82, CI: 0.72-0.94, *p<0.001*), spending (AOR=0.61, CI: 0.53-0.71, *p<0.000*), visiting families (AOR=0.79, CI: 0.70-0.88, *p<0.000*), and not using contraception (AOR=0.58, CI: 0.42-0.80, *p<0.000*) were significantly associated with decreased experience of emotional violence.

**Table 2:** Multiple logistic regression models indicating unadjusted and adjusted odds ratios with [95% confidence intervals] for the association between types of violence and categories of women’s autonomy (WA) among Afghan married Women ages 15-49, AfDHS 2015.

|  | M1: Unadjusted | M2: M1+sociodem. | M3: M2+all WA |
| --- | --- | --- | --- |
| **Physical Violence** |  |  |  |
| Healthcare | 0.65 [0.5-0.76]*** | 0.70 [0.60-0.81]*** | 1.02 [0.88-1.19] |
| Spending | 0.59 [0.51-0.67]*** | 0.58 [0.51-0.66]*** | 0.71 [0.61-0.82]*** |
| Household purchase | 0.59 [0.51-0.68]*** | 0.59 [0.52-0.68]*** | 0.72 [0.61-0.84]*** |
| Visiting family | 0.68 [0.57-0.78]*** | 0.69 [0.60-0.80]*** | 0.97 [0.83-1.14] |
| Contraception | 0.92 [0.64-1.33] | 0.97 [0.68-1.38] | 1.01 [0.69-1.47] |
| Not using | 0.68 [0.47-0.97]* | 0.66 [0.46-0.93]* | 0.70 [0.48-1.02] |
| **Sexual Violence** |  |  |  |
| Healthcare | 0.54 [0.41-0.70]*** | 0.51 [0.39-0.65]*** | 0.55 [0.41-0.73]*** |
| Spending | 0.71 [0.54-0.92]* | 0.62 [0.48-0.80]*** | 0.99 [0.76-1.30] |
| Household purchase | 0.63 [0.49-0.80]*** | 0.56 [0.43-0.72]*** | 0.69 [0.49-0.98]* |
| Visiting family | 0.86 [0.68-1.08] | 0.78 [0.63-0.97]* | 1.29 [0.94-1.77] |
| Contraception | 1.07 [0.61-1.86] | 1.14 [0.65-1.99] | 1.22 [0.69-2.15] |
| Not using | 0.91 [0.53-1.55] | 0.98 [0.58-1.67] | 0.50 [0.61-1.80] |
| **Emotional Violence** |  |  |  |
| Healthcare | 0.78 [0.68-0.89]** | 0.82 [0.72-0.94]** | 1.15 [0.95-1.39] |
| Spending | 0.64 [0.55-0.74]*** | 0.61 [0.53-0.71]*** | 0.62 [0.50-0.78]*** |
| Household purchase | 0.75 [0.61-0.92]** | 0.74 [0.60-0.91]** | 0.89 [0.62-1.29] |
| Visiting family | 0.77 [0.68-0.87]*** | 0.79 [0.70-0.88]*** | 0.95 [0.77-1.17] |
| Contraception | 0.86 [0.61-1.21] | 0.92 [0.66-1.29] | 0.95 [0.67-1.35] |
| Not using | 0.53 [0.38-0.74]*** | 0.58 [0.42-0.80]** | 0.60 [0.42-0.85]** |

*Footnotes:* Significance: <0.05\*, 0.00\*\*, 0.000\*\*\*
M1: unadjusted odds ratios [95% confidence intervals]
M2: adjusted for age, residency, wealth, parity, and ethnicity
M3: adjusted for all the covariates including the five WA variables (decision making in healthcare, spending, household purchases, visiting family, and contraception), age, residency, wealth, parity, and ethnicity

In Model 3, in addition to adjusting for the confounders, we adjust for the five women’s autonomy variables (decision making in healthcare, spending, household purchases, visiting family, and contraception) We present Model 3 because we found small multicollinearity (VIF of 6) only with the autonomy in the use of contraception variable. Multicollinearity for all other autonomy variables was less than VIF of 3. In Model 3, we found that WA in spending (AOR=0.71, CI: 0.61-0.82, *p<0.000*) and household purchases (AOR=0.72, CI: 0.61-0.84, *p<0.000*) remained associated with physical violence, while WA in healthcare (AOR=0.55, CI: 0.41-0.73, *p<0.000*) and household purchases (AOR=0.69, CI: 0.49-0.98, *p<0.01*) remained associated with sexual violence. Lastly, WA in spending (AOR=0.62, CI: 0.50-0.78, *p<0.000*) and not using contraception (AOR=0.60, CI: 0.42-0.85, *p<0.001*) remained associated with emotional violence.

**Table 3** shows the association between categories of women’s autonomy and domestic violence types stratified by education (any vs. none). After adjusting for all covariates including women’s autonomy variables, we found that there was a greater protective effect of WA in visiting family among women with any education (vs. no education) across each domestic violence outcome--physical violence: (AOR=0.44 vs. AOR=0.75, *p<0.001*), sexual violence: (AOR=0.39 vs. AOR=0.86, *p<0.039*), and emotional violence: (AOR=0.43 vs. AOR=0.88, *p<0.000*). There was no evidence of interaction between education and other WA variables.

**Table 3:**
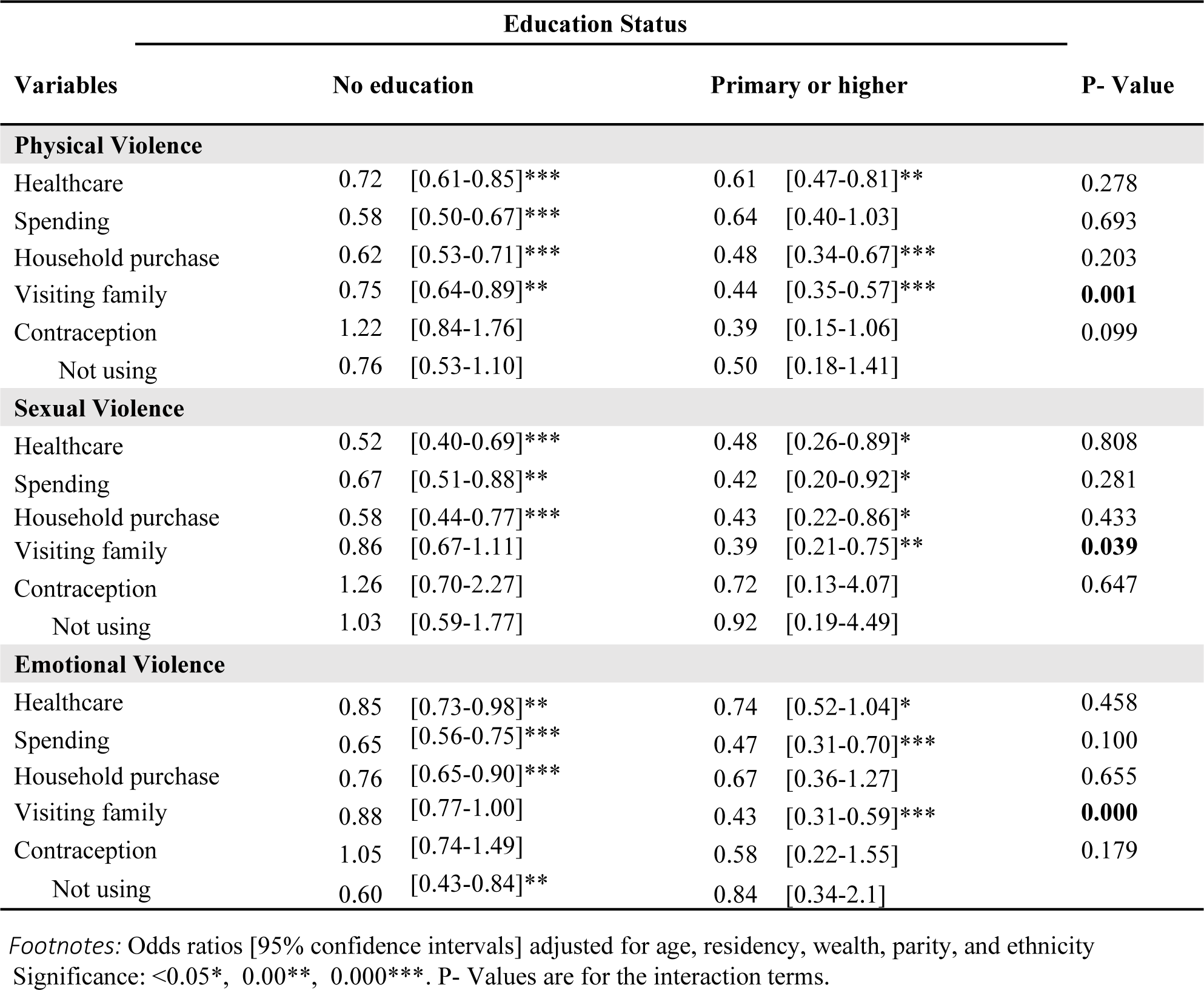
Interaction of education as a moderator of the association between categories of women’s autonomy and domestic violence types among Afghan married Women ages 15-49, AfDHS 2015.

| Variables | Education Status |  |  |  | P- Value |
| --- | --- | --- | --- | --- | --- |
|  | No education |  | Primary or higher |  |  |
| Physical Violence |  |  |  |  |  |
| Healthcare | 0.72 | [0.61-0.85]*** | 0.61 | [0.47-0.81]** | 0.278 |
| Spending | 0.58 | [0.50-0.67]*** | 0.64 | [0.40-1.03] | 0.693 |
| Household purchase | 0.62 | [0.53-0.71]*** | 0.48 | [0.34-0.67]*** | 0.203 |
| Visiting family | 0.75 | [0.64-0.89]** | 0.44 | [0.35-0.57]*** | **0.001** |
| Contraception | 1.22 | [0.84-1.76] | 0.39 | [0.15-1.06] | 0.099 |
| Not using | 0.76 | [0.53-1.10] | 0.50 | [0.18-1.41] |  |
| Sexual Violence |  |  |  |  |  |
| Healthcare | 0.52 | [0.40-0.69]*** | 0.48 | [0.26-0.89]* | 0.808 |
| Spending | 0.67 | [0.51-0.88]** | 0.42 | [0.20-0.92]* | 0.281 |
| Household purchase | 0.58 | [0.44-0.77]*** | 0.43 | [0.22-0.86]* | 0.433 |
| Visiting family | 0.86 | [0.67-1.11] | 0.39 | [0.21-0.75]** | **0.039** |
| Contraception | 1.26 | [0.70-2.27] | 0.72 | [0.13-4.07] | 0.647 |
| Not using | 1.03 | [0.59-1.77] | 0.92 | [0.19-4.49] |  |
| Emotional Violence |  |  |  |  |  |
| Healthcare | 0.85 | [0.73-0.98]** | 0.74 | [0.52-1.04]* | 0.458 |
| Spending | 0.65 | [0.56-0.75]*** | 0.47 | [0.31-0.70]*** | 0.100 |
| Household purchase | 0.76 | [0.65-0.90]*** | 0.67 | [0.36-1.27] | 0.655 |
| Visiting family | 0.88 | [0.77-1.00] | 0.43 | [0.31-0.59]*** | **0.000** |
| Contraception | 1.05 | [0.74-1.49] | 0.58 | [0.22-1.55] | 0.179 |
| Not using | 0.60 | [0.43-0.84]** | 0.84 | [0.34-2.1] |  |

Footnotes: Odds ratios [95% confidence intervals] adjusted for age, residency, wealth, parity, and ethnicity Significance: <0.05\*, 0.00\*\*, 0.000\*\*\*. P- Values are for the interaction terms.

## Discussion

In this context of a high prevalence of violence and low autonomy, we found that about one in two Afghan women did not have autonomy in making decisions about spending, household purchases, and visiting family, and more than half of the sample did not have autonomy in making decisions about their healthcare and use of contraception. Additionally, almost one in two women experienced physical violence, and one in three experienced emotional violence. After adjustment for sociodemographic factors, almost all associations with women’s autonomy were significant (with the exception of contraception). Even when women’s autonomy variables were added together in the model, a few women’s autonomy domains still remained significant. The factors that remained significant were related to healthcare as well as spending and household purchases. A potential explanation is that women, who lack financial autonomy and are dependent on their partner, may not have the option to leave abusive relationships, and therefore, continue to experience physical and/or sexual violence.^18^ Our results are consistent with earlier investigations carried out in Bangladesh^18^ and Ghana,^11^ which found a significant association between women’s autonomy and decreased experience of physical and sexual violence.

We also found that women who had the autonomy to visit families were less likely to experience physical, sexual, or emotional violence if they had at least primary education (vs. no education). An explanation for these findings is might be that educated women have a better ability to navigate travel by themselves and negotiate decision making with their partners. Previous studies show that, compared to women with no education, women with some education are more likely to have decision making autonomy^37,38^ and less likely to experience physical or emotional violence.^11^ Unfortunately, the 2015 demographic health survey shows that 86% of women have no education,^4^ and this number increases with the recent ban on women’s education by the Taliban.^34,35^

We are unaware of other studies that have assessed the impact of Afghan women’s lack of autonomy on their experience of domestic violence and the moderating effect of education on this association. Since our study sample is from a nationally representative dataset, our findings are generalizable to Afghanistan. Furthermore, we looked at multiple components of WA and multiple types of violence, whereas, other studies have grouped them together – this allows an understanding of types of autonomy and how each type of WA might remain associated with violence. Additionally, we looked at domestic violence, rather than just intimate partner violence, as the contextual factors around WA may affect violence more broadly in the household or through in-laws and relatives rather than just a partner.

Our study also has some limitations, including the potential for social desirability bias regarding reporting of domestic violence. This bias was somewhat minimized by asking women about domestic violence privately in the absence of their husbands in order for women to feel safe and give accurate information. In addition, concerns over temporality may be an issue in cross-sectional studies. In this study it is likely that reporting on women’s autonomy reflects experiences throughout the course of the relationship. Additionally, we restricted the outcome to domestic violence experienced in the past 12 months to ensure the outcome occurred recently relative to the interview period to better ensure the exposure preceded the outcome. Although the data was collected in 2015 and there have been changes in the Afghan government since then, our discoveries are likely still pertinent because the circumstances of women in the country have not improved and have, likely, gotten worse in the country in recent years.^36^

This study will provide evidence and bring awareness that is needed to urge policymakers and program implementers in targeting and improving women’s autonomy in Afghanistan. Considering the current women’s condition under the Taliban administration, this research has significant relevance. Beyond filling the current gap in the existing public health literature, this has implications for understanding the potential adverse outcomes that may be exacerbated by the Taliban party’s policies, including eliminating the Ministry of Women’s Affairs, suspending women’s education, and banning women’s freedom of movement in public unless they are escorted by a male relative.^34,36^ Without autonomy and education, women may be more susceptible to experiencing domestic violence. This not only has an impact on the health of women but has also been linked to other reproductive and perinatal health outcomes^16–25^ and could have generational implications. Future studies should assess the impact of women’s autonomy on birth outcomes (e.g., pregnancy loss) and how this association may be mediated by experience of domestic physical violence, since a previous study in Afghanistan shows domestic physical violence to be associated with pregnancy loss.^42^

## Data Availability

We used Afghanistan 2015 DHS publicly available data

## References

1. Dyson T, Moore M. On Kinship Structure, Female Autonomy, and Demographic Behavior in India. Popul Dev Rev. 1983;9(1):35. doi:10.2307/1972894

2. Safilios-Rothschild C. Female power, autonomy and demographic change in the Third World. In: Anker R, M B, Youssef NH, eds. Women’s Roles and Population Trends in the Third World. Croon Helm; 1982:117–132.

3. In focus: Sustainable Development Goal 5. UN Women – Headquarters. Accessed December 28, 2022. https://www.unwomen.org/en/news-stories/in-focus/2022/08/in-focus-sustainable-development-goal-5

4. Central Statistics Organization; Ministry of Public Health (CSO & MoPH. Published online 2017. https://www.dhsprogram.com/pubs/pdf/FR323/FR323.pdf

5. Dixon RB. Rural Women at Work: Strategies for Development. RFF Press; 2013.

6. Bhandari TR, Kutty VR, Ravindran TK. Women’s Autonomy and Its Correlates in Western Nepal: A Demographic Study. PLoS One. 2016;2016;11(1):e0147473.

7. Ristiana R, Handayani D. Does work influence women’s autonomy or does autonomy deliberate women to work? Herdiansyah H, ed. E3S Web Conf. 2018;74:10013. doi:10.1051/e3sconf/20187410013

8. Mistry R, Galal O, Lu M. Women’s autonomy and pregnancy care in rural India: a contextual analysis”. Soc Sci Med. 2009;69(6):926–933.

9. Kc S, Neupane S. Women’s Autonomy and Skilled Attendance During Pregnancy and Delivery in Nepal. Matern Child Health J. 2016;20(6):1222–1229.

10. Ahmed F, Oni FA, Hossen SS. Does gender inequality matter for access to and utilization of maternal healthcare services in Bangladesh? PLoS One. 2021;16(9).

11. Tenkorang EY. Women’s autonomy and intimate partner violence in Ghana. Int Perspect Sex Reprod Health. 2018;1;44(2):51–61.

12. Bauserman M, Lokangaka A, Thorsten V. Risk factors for maternal death and trends in maternal mortality in low- and middle-income countries: a prospective longitudinal cohort analysis. Reprod Health. 2015;12(Suppl 2).

13. Goal 5 | Department of Economic and Social Affairs. Accessed June 24, 2023. https://sdgs.un.org/goals/goal5

14. Shannon G, Jansen M, Williams K, et al. Gender equality in science, medicine, and global health: where are we at and why does it matter? The Lancet. 2019;393(10171):560–569. doi:10.1016/S0140-6736(18)33135-0

15. Health - United Nations Sustainable Development. Accessed June 24, 2023. https://www.un.org/sustainabledevelopment/health/

16. Rahman MM, Mostofa MG, Hoque MA. Women’s household decision-making autonomy and contraceptive behavior among Bangladeshi women. Sex Reprod Healthc. 2014;1;5(1):9–15.

17. Sano Y, Antabe R, Atuoye KN, Braimah JA, Galaa SZ, Luginaah I. Married women’s autonomy and post-delivery modern contraceptive use in the Democratic Republic of Congo. BMC Womens Health. 2018;18(1).

18. Rahman M, Nakamura K, Seino K, Kizuki M. Does gender inequity increase the risk of intimate partner violence among women? Evidence from a national Bangladeshi sample [published correction appears in PLoS One. 2014;9(2):e91448. PLoS One. 2013;8(12).

19. Abada T, Tenkorang EY. Women’s autonomy and unintended pregnancies in the Philippines. J Biosoc Sci. 2012;44(6):703–718.

20. Besera G, Roess A. The relationship between female genital cutting and women’s autonomy in Eritrea. Int J Gynaecol Obstet. 2014;126(3):235–239.

21. Memiah P, Opanga Y, Bond T. Is sexual autonomy a protective factor for neonatal, child, and infant mortality? A multi-country analysis. PLoS One. 2019;14(2).

22. Chakraborty P, Anderson AK. Maternal autonomy and low birth weight in India. J Womens Health. 2011;1;20(9):1373–82.

23. Sharma A, Kader M. Effect of women’s decision-making autonomy on infant’s birth weight in rural Bangladesh. Int Sch Res Not. 2013;2013.

24. Klugman J, Li L, Barker KM, Parsons J, Dale K. How are the domains of women’s inclusion, justice, and security associated with maternal and infant mortality across countries? Insights from the Women, Peace, and Security Index [published correction appears in SSM Popul Health. 2020 Dec 10;12:100716. SSM Popul Health. 2019;9(100486).

25. Adhikari R, Sawangdee Y. Influence of women’s autonomy on infant mortality in Nepal. Reprod Health. 2011;8(7).

26. Hindin M, Adair L. Who’s at risk? Factors associated with intimate partner violence in the Philippines. Soc Sci Med. 2002;55(8):0277-9536 01 00273-8 18. doi:doi:10.1016/

27. Paul S. Women’s Labour Force Participation and Domestic Violence: Evidence from India. J South Asian Dev. 2016;11(2):224–250. doi:10.1177/0973174116649148

28. Pallitto CC, O’Campo P. Community level effects of gender inequality on intimate partner violence and unintended pregnancy in Colombia: testing the feminist perspective. Soc Sci Med. 2005;60(10):2205–2216.

29. Jejeebhoy SJ, Sathar Z. Women’s Autonomy in India and Pakistan: The Influence of Religion and Region. Popul Dev Rev. 2001;27:687–712.

30. Runion ML. The History of Afghanistan. ABC-CLIO; 2017.

31. Quesada J, Hart LK, Bourgois P. Structural vulnerability and health: Latino migrant laborers in the United States. Med Anthropol. 2011;30(4):339–362.

32. Jewkes R, Corboz J, Gibbs A. Trauma exposure and IPV experienced by Afghan women: analysis of the baseline of a randomised controlled trial. PLoS One. 2018;13(10):0201974.

33. Organization WH. Addressing Violence against Women in Afghanistan: The Health System Response (No. WHO/RHR/15.26). World Health Organization; 2015.

34. Mackintosh EC. Taliban decree on women’s rights makes no mention of school or work. Published online December 4, 2021. https://edition.cnn.com/2021/12/03/asia/afghanistan-taliban-decree-womens-rights-intl/index.html

35. The 10 Best and Worst Countries to Be a Woman in 2021. Accessed December 28, 2022. https://www.globalcitizen.org/en/content/best-worst-countries-for-women-gender-equality/

36. Kottasová EP Ivana. Taliban suspend university education for women in Afghanistan. CNN. Published December 20, 2022. Accessed December 28, 2022. https://www.cnn.com/2022/12/20/asia/taliban-bans-women-university-education-intl/index.html

37. Haque SE, Rahman M, Mostofa MG, Zahan MS. Reproductive health care utilization among young mothers in Bangladesh: does autonomy matter? Womens Health Issues. 2012;22(2).

38. Nigatu D, Gebremariam A, Abera M, Setegn T, Deribe K. Factors associated with women’s autonomy regarding maternal and child health care utilization in Bale Zone: a community based cross-sectional study. BMC Womens Health. 2014;14(79).

39. Ahmadi R, Soleimani R, Jalali MM, Yousefnezhad A, Rad M, Eskandari A. Association of intimate partner violence with sociodemographic factors in married women: a population-based study in Iran Association of intimate partner violence with sociodemographic factors in married women: a population-based study in Iran. Published online September 30, 2016.

40. The DHS Program - Quality information to plan, monitor and improve population, health, and nutrition programs. Accessed December 29, 2022. https://dhsprogram.com/

41. Multicollinearity in Regression Analyses Conducted in Epidemiologic Studies - PMC. Accessed November 9, 2023. https://www.ncbi.nlm.nih.gov/pmc/articles/PMC4888898/

42. Ibrahimi S, Alamdar Yazdi A, Yusuf KK, Salihu HM. Association of domestic physical violence with feto-infant outcomes in Afghanistan. Asia Pac J Public Health. 2021;Mar;33(2-3):273–9.

